# Impact of Dobbs v. Jackson on Abortion Access in Colorado: An Analysis of Incidence and Demographic Shifts Post-Roe

**DOI:** 10.1101/2025.05.21.25328096

**Authors:** Abigail Bryer, Thomas McAndrew, Fathima Wakeel, Christine Daley

## Abstract

**Objectives:** To estimate the impact in the number of incident abortions due to the Dobbs v Jackson decision.

**Methods:** We fit an interrupted time series model to annual incident abortions from 2004 to 2023 in the state of Colorado. In addition, we computed the proportion of residents that received abortive care from outside Colorado before and after the Dobbs v Jackson decision. Finally, we estimated the proportion of abortions over time stratified by gestation week of the mother.

**Results:** We found that in the state of Colorado, after Dobbs v Jackson (compared to before): (1) an increase in non-resident incident abortions immediately after the decision (2) which is driven by residents in the states surrounding Colorado with the most restrictive abortion laws; the uptick in non-resident abortions has likely led to longer wait times, a higher burden on facilities, and (3) later gestation times for the mother.

**Conclusions:** Shifts in abortion patterns in Colorado is evidence that the ramifications of Dobbs v Jackson extend beyond individual state borders, impacting both state-residents and non-residents seeking care. Policymakers must consider these findings in future reproductive legislation to ensure access to abortion in protective states, such as Colorado.

**Public health implications:** This work underscores the immediate impact of healthcare legislation and the increased burden on healthcare services for states with more permissible abortion legislation.

## Introduction

Dobbs v Jackson (DvJ) was ratified on June 24, 2022, overturning Roe v Wade (RvW), which provided federal abortion protection in the United States from 1973 until 2022. The Dobbs decision returned the power to regulate abortion to individual states through re-interpretation of the fourteenth amendment of the US Constitution as not conferring a right to an abortion. Since the 1976 Hyde Amendment, federal funding of abortions has not been allowed; the three exceptions to this, life endangerment of the mother, rape, and incest, were approved in 1994.^1^ Based on the Dobbs decision, no federal funding of abortions can occur, regardless of the circumstances. Thus, states now have the power to both regulate and determine public funding of abortions; at the time of the writing of this article, 14 states have a total abortion ban and 27 have partial bans based on gestational duration. As the number of state-level abortion restrictions increase, it is estimated to cost state economies in those states without a ban more than $105 billion dollars per year, with $1.2 billion dedicated to Medicaid costs for unplanned births.^2, 3^

Preliminary evidence suggests unanticipated sequelae of Dobbs include increased maternal: (1) mortality, (2) comorbidities associated with mortality, and (3) mental health conditions. It also disproportionately affects individuals with decreased socioeconomic status. It is estimated that abortion bans across the country will result in an increase of 21% in maternal mortality on average, with higher predicted prevalence for racial and ethnic minorities.^5^ Women denied abortion are more likely to experience gestational hypertension, which has the potential to lead to eclampsia.^6^ Gestational hypertension has also been found to be associated with higher likelihood of recurrence in subsequent pregnancies.^6^ In addition to bodily harm, women denied abortion are more likely to suffer from anxiety, depression, and low self-esteem in the short-term period after being denied care.^6^ Compared to women who are able to access abortion care, women who cannot access abortion have a 4 times greater odds of living below the Federal Poverty Level.^6^

In addition to the health burdens on the individual, clinics that serve women and public health services have experienced higher abortion incidence for non-resident patients, longer appointment waiting times, extended hours, and more anti-abortion sentiment.^7, 8^ The immediate effects of DvJ resulted in longer wait times (up to three weeks) in states that protect the right to abortion and are adjacent to states that have banned abortion.^9^ Since the initial peak in the months following June 24, 2022, wait times for abortion have declined but still remain longer than when RvW was in place.^7^ Abortion facilities are (1) at risk of being overwhelmed by appointment demand and unable to treat all patients, as well as (2) being subject to more frequent anti-abortion violence. Abortion clinics in 2022 saw increases in bomb threats by 133%, obstructions of clinic entrances by 538%, assault and batteries by 29%, and burglaries by 100%.^8^ The delays experienced in abortion facilities have created an estimated demand for 4,300-16,500 late term abortions nationwide.^10^

In this study, we hypothesized that states that have protected the right to an abortion have experienced higher volumes of abortion incidence compared to average annual rates since the Dobbs decision, and that this influx was driven by non-resident patients. We used data from Colorado to support this claim, showing the observed changes in total and non-resident incidence in recent years. This work shows the significance of changes driven by non-resident patients coming from states that are restrictive or have banned abortion and begins to address our limited understanding of the effects DvJ has on states with protective legislation.

## Methods

### Data Collection

We collected state level numbers of abortions performed at a health care facility for Colorado from 1967 to 2023 via the Colorado Department of Public Health and Environment (CDPHE). The CDPHE mandates abortion reporting to the state and collects counts at a yearly and monthly cadence. Abortion reports are inclusive of terminations occurring at physician clinics and hospitals. De-identified data were collected from the CDPHE via a data request form and the Lehigh University Human Subjects Committee determined that no Institutional Review Board approval was necessary for this study.

The following demographic data about women were available for Colorado: (1) race/ethnicity, (2) gestation week at time of termination, (3) age, (4) resident or non-resident status, (5) marital status, and (6) type of procedure (medical nonsurgical, suction curettage, dilation and evacuation, sharp curettage, intrauterine instillation, hysterectomy, or other). We appended (7) a binary indicator variable that takes the value 0 before the Dobbs v Jackson decision and the value 1 on and after Dobbs v Jackson. From 2020 to 2023, the CDPHE collected and reported the state of residence for each abortion patient; the provided data represented missing states as having < 4 residents travel for abortion.

### Analysis

Continuous variables were summarized as mean and standard deviation. Categorical variables were summarized as percent and frequency. The estimated mean incident number of abortions before and after Dobbs v Jackson was assessed with a two-sample Student’s t-test. The chi-square test was used to test for an association between categorical variables. A p-value smaller than 0.05 was considered statistically significant.

We fit an Interrupted Time Series (ITS) model to evaluate the change in abortion incidence stratified by total, non-resident, and resident abortions. Our model assumes the expected number of abortions (E(*Y*)) depends on the elapsed number of years since 2004 (*T*), an indicator variable that equals 1 on and after Dobbs v Jackson and equals 0 before this event (*D*), and the elapsed number of months after the DvJ decision (*P*); E(*Y*) = (*b0 +b2D) + (b1T+b3P)*, where *bx* are model parameters. We assume residuals are Normally distributed and fit this model using ordinary least squares regression.

To explore the difference in the number of abortions from patients outside Colorado, we generated choropleth maps. Evidence for an association between changes in the number of abortions from non-resident patients from 2020-2022 was tested via Chi-square.

## Results

### Demographics

After Dobbs V Jackson we observed an increase in non-resident abortions (before DvJ: 10.94%, after DvJ: 31.82%; p <0.01), a suspected increase in the proportion of medical non-surgical abortions (before: 42.82%; after: 69.16%; p = 0.10), and an increase in the proportion of abortions after gestation week 23 (before: 0.15%; after: 0.75%, p < 0.01). See Table 1.

**Table 1.**
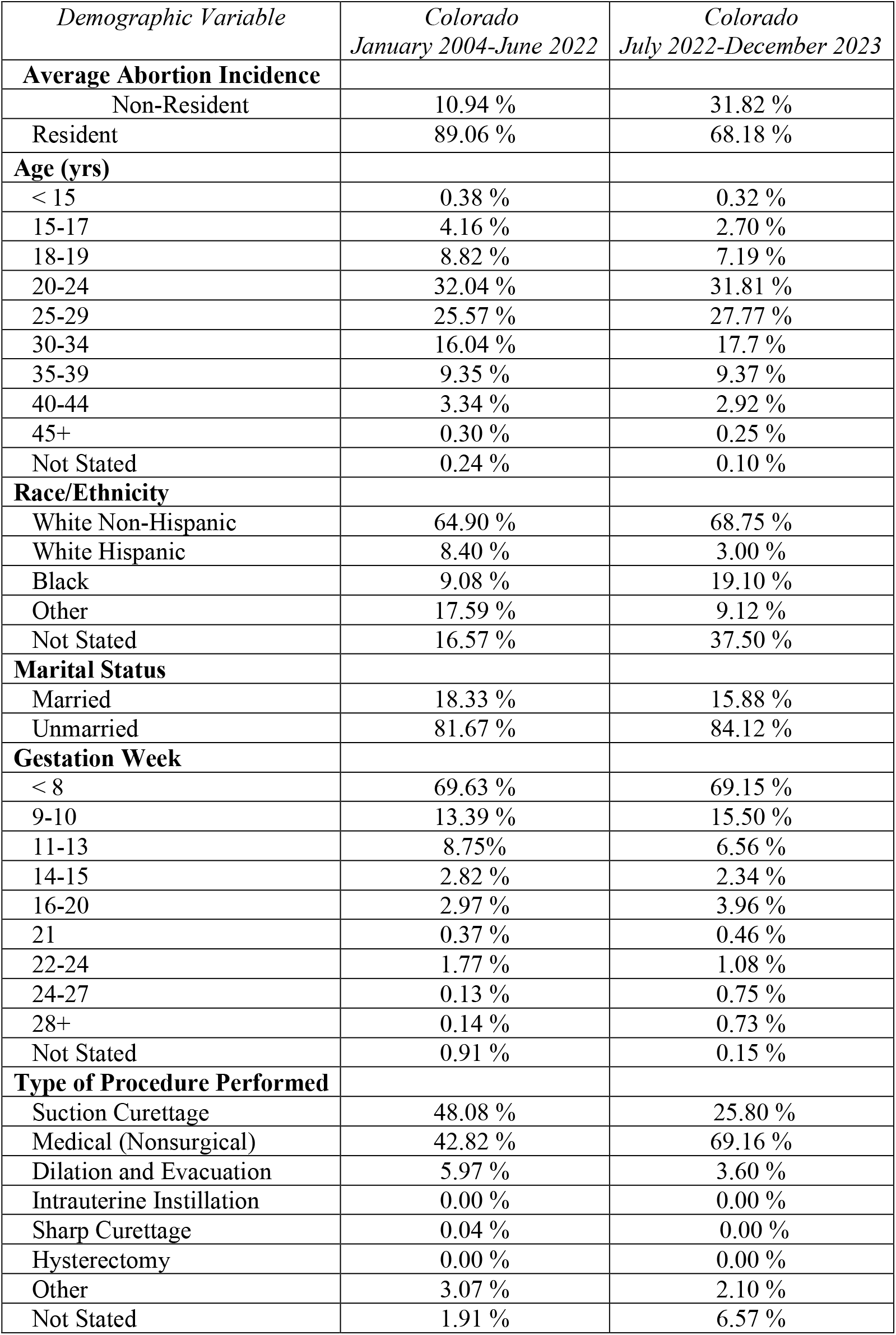
Summary of monthly incident abortions and associated demographic data for Colorado stratified by (left column) those months before the Dobbs vs Jackson decision (Jan. 2004 to June 2022) and (right column) those months after the decision (July 2022 to Dec. 2023).

### Increase in non-resident abortion incidence after Dobbs v Jackson decision

The total number of monthly incident abortions in Colorado increased after DvJ (before 878; after 1180; p<0.01). This number, however, does not provide the full picture. We observed a 2.89 times increase in the number of non-resident abortions after DvJ (monthly mean incident cases = 366) compared to before DvJ (monthly mean incident cases = 94); p<0.01; Table 2), while the number of incident abortions among residents increased only 5.6% (p=0.19) from a mean of 782 to 818 abortions.

**Table 2.** The mean total, non-resident, and resident monthly incident abortions before (left column) and after (right column) the Dobbs vs Jackson decision, the relative difference, and pvalue to test if there was a significant difference.

| *Population* | *Before DvJ (January 2004-June 2022)* | *After DvJ (July 2022-December 2023)* | *Relative difference* | *Two-sample pvalue* |
| --- | --- | --- | --- | --- |
| **Total Abortion Incidence** | 878 | 1190 | 35.5% | <0.01 |
| **Non-resident Abortion Incidence** | 94 | 366 | 289.3% | <0.01 |
| **Resident Abortion Incidence** | 782 | 818 | 5.6% | 0.19 |

**Table 3.** Colorado annual interrupted time series analysis statistics.

| Population | Point estimate | 95% CI | p-value |
| --- | --- | --- | --- |
| Annual Total Abortion Incidence 2004-2023 |  |  |  |
| Change in rate of incident abortions after DvJ (*P*) | 49.65 | [-162.008, 261.304] | 0.63 |
| Shift in abortion incidence at time of DvJ (*D*) | 4391.56 | [491.119, 8292.00] | 0.03 |
| Annual Non-resident Abortion Incidence 2004-2023 |  |  |  |
| Change in rate of incident abortions after DvJ (*P*) | 25.45 | [-13.696, 64.604] | 0.19 |
| Shift in abortion incidence at time of DvJ (*D*) | 2449.66 | [1728.189, 3171.128] | <0.01 |
| Annual Resident Abortion Incidence 2004-2023 |  |  |  |
| Change in rate of incident abortions after DvJ (*P*) | 17.61 | [-170.227, 205.448] | 0.85 |
| Shift in abortion incidence at time of DvJ (*D*) | 2014.32 | [-1447.186, 5475.822] | 0.24 |

### Marked increase in incident abortions near the Dobbs v Jackson decision in time

We found a 2,449 (95CI = [1,728, 3,171]; p <0.01) increase in the number of non-resident abortions and 4,391 increase in total (resident plus non-resident) number of abortions (95CI = [491, 8,292]; p = 0.03) in the six months, and year after, that DvJ was ratified (See table 2 for Interrupted Time Series analysis estimates and Fig. 1). There was an approximate increase of 2,014 resident abortions (95CI = [-1447, 5475]) during this time period (p = 0.24). An increase in the rate of incident abortions after the DvJ was not statistically significant, but the maximum likelihood estimate suggested a positive increase.

**Figure 1.**
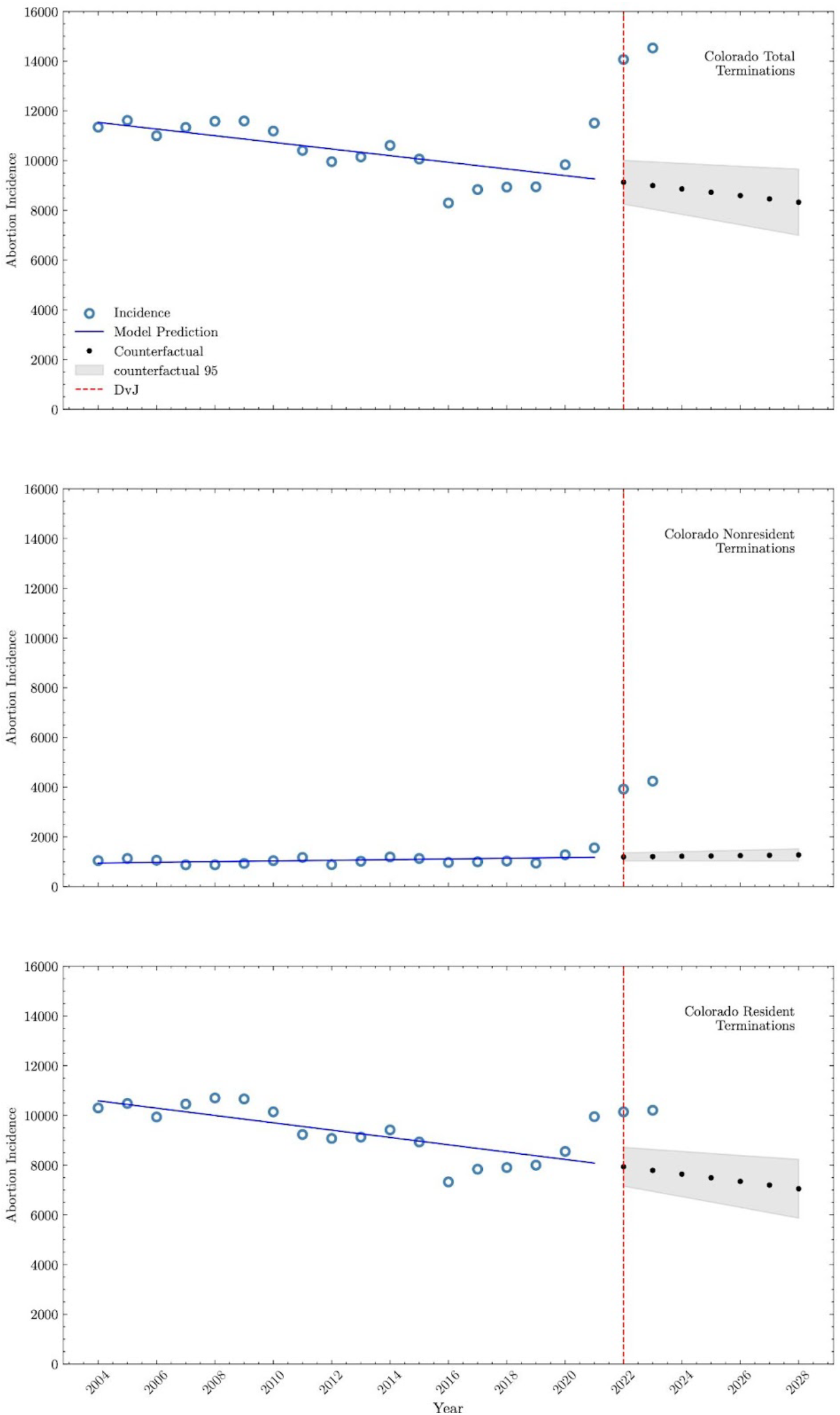
Interrupted time series analysis for Colorado total terminations, non-resident terminations, and resident terminations from 2004-2023, including long-term trend projections.

### Surrounding states that experienced changes in the rate of non-resident abortions to Colorado

Figure 2 depicts the proportion of non-resident abortions from each state for the years 2020 (n = 1242.0) to 2023 (n = 4142.0). Of all states, Texas had a 277.78% increase in the proportion of residents receiving abortion care in Colorado from 2020 (0.18%) to 2023 (0.68%). In Colorado, the mean incident cases of Texas residents grew from 316.5 before DvJ to 2632.0 after DvJ, a 731.6% increase. Colorado also experienced a significant change in the proportion of Oklahoma state residents from 0.01% to 0.04% of the total non-resident abortions, an increase of 300%. Total Oklahoma abortions changed from a mean incidence of 13.5 before 2022 to a mean incidence of 189.5, an increase of 1303.7%.

**Figure 2.**
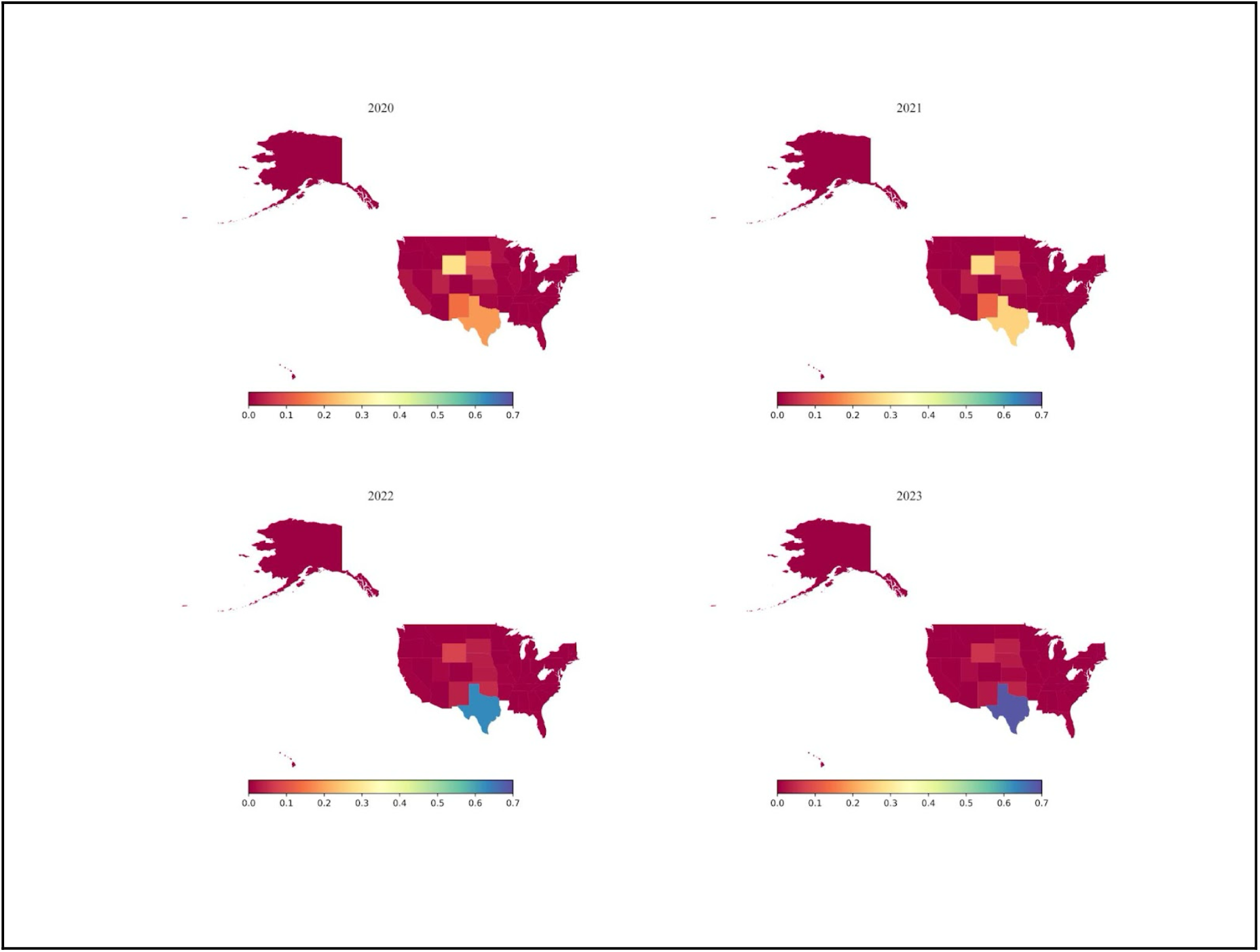
Choropleths tracking the proportion of state residence of non-resident abortions performed in Colorado from 2020-2023.

### Average gestation week during termination is later after DvJ decision

After the DvJ decision, the average number of monthly abortions increased for mothers with later gestation weeks (see Fig. 3): (1) 9-13 weeks increased from 1,923 on average in 2020-2021 to 2,945 on average in 2022-2023 or a 53.14% increase; (2) 14-24 weeks increased from 743 on average in 2020-2021 to 1,185 on average in 2022-2023 or a 59.48% increase; and, most notably, (3) 25-28+ weeks increased from 32 average in 2020-2021 to 235 average in 2022-2023 or a 634.37% increase. In comparison, from 2004 to 2021, the average number of abortions occurring in gestation weeks 25-28+ equaled 15 per year.

**Figure 3.**
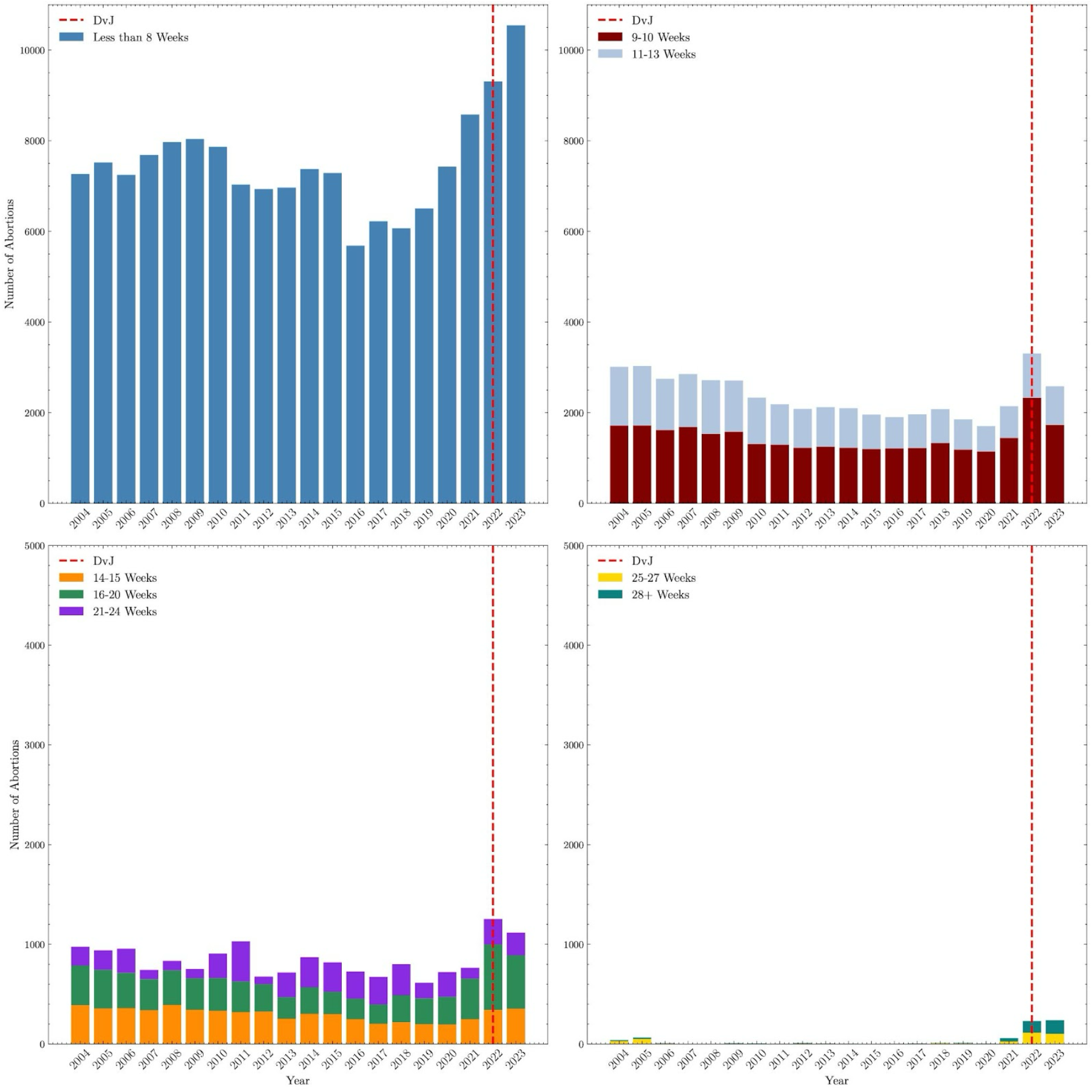
Prevalence of abortion procedures stratified by gestation week in Colorado from 2004-2023.

## Discussion

Data collected from Colorado on incident abortions from 2004 to the present suggest that the Dobbs decision has resulted in an increase in total terminations driven by non-residents from the states surrounding Colorado. The uptick in non-resident abortions has likely led to longer wait times, a higher burden on facilities, and later gestation times for the mother.

Restrictive abortion legislation enacted in individual states following DvJ has led to residents traveling to states with less restrictive laws for care. The ratification of DvJ resulted in 13 states’ trigger laws severely restricting or banning abortion almost immediately; currently there are 14 total state bans and 27 partial bans as of June 2024. Of these states, Texas and Oklahoma have banned abortion with very limited exceptions.^11^. Colorado has since experienced tremendous increases in non-resident abortions from surrounding states like Texas and Oklahoma (see figure 2). Prior to 2022, Texas reported on average 50,000 in-state abortions annually, and only 62 in 2023. ^12^ In 2023 Oklahoma reported a total of 0 in-state abortions compared to an average 5,000 before DvJ.^13, 14^ Our data suggest restrictive states like Texas and Oklahoma are displacing residents seeking abortion onto states with less restrictive laws, such as Colorado. Additional examination of incident cases in other states with greater protections for those seeking abortions is warranted to better understand what is happening at a national level.

The immediate increase in the number of out-of-state residents seeking care after DvJ has resulted in longer wait times for patients. Planned Parenthood of the Rocky Mountains has reported wait times of 28 days immediately after DvJ, 11 days longer than the typical wait time of 17 days in the months prior to the decision.^15^ The 17-day wait times in Colorado likely were a result of the enacting of the Texas Heartbeat Act and the creation of Texas’s trigger law in 2021. Otherwise in the years leading up to DvJ, many states had average wait times of only 5 days.^15^

Use of telehealth in Colorado and similar states has been important in improving appointment wait times for prescriptions of medical nonsurgical abortions to patients living in restrictive states.^16^ Telehealth appointments that can prescribe medical abortions using medications such as Mifepristone and Misoprostol for women with a gestation time of less than 10 weeks have increased in response to excess burden at healthcare facilities. In Colorado, before DvJ, the proportion of medical terminations was 42.82% and after, 69.16%. Though telehealth may be a way for some individuals to receive abortion care in a timely fashion, it is only useful for early abortions and, thus, does not take the place of surgical care for many women.

Appointment delays in Colorado may be a driving factor behind the 634.37% increase in abortions after gestation week 25 that was observed in 2022-2023. Colorado has reported experiencing higher volumes of patients seeking abortion in later stages of pregnancy due to the several weeks of delays they are facing. Colorado is also one of only 8 states with abortion legislation that does not restrict termination at any gestation week.^11, 15^ Unfortunately for patients, as appointment delays push them later into pregnancy, the cost for an abortion increases from an average $800 in the first trimester to an average cost between $1,500-$2,000 in the second trimester.^17^ Maternal health concerns also increase as abortion is pushed further into the later stages of pregnancy; being denied abortion is associated with higher risk of eclampsia, postpartum hemorrhage, gestational hypertension, and a four times greater odds of living below the federal poverty line.6

The shift in gestational age at time of termination to later in pregnancy complicates the impact of the Dobbs decision. Many states enacted restrictions on abortions, not complete bans. These restrictions were based primarily on gestation time. The influx of patients seeking care from states with bans on abortions has caused greater waiting periods, likely including in states that have gestational restrictions. Thus, people seeking abortions in these states may miss the time period that is legal in their state, sending them to states like Colorado without limitations. As a result, Colorado and other states with similar laws will likely face even more increases in late-term abortions with higher cost and greater health concerns.

In addition to financial costs changing at an individual level, the high costs of abortions impact health care spending at the state and national levels. National reproductive organizations have had to increase their spending to assist patients in restrictive states. Since DvJ, Planned Parenthood has increased its patient assistance funding from $1.2 million in the year prior to DvJ to over $9 million in the year after.^18^ Between 2021 and 2022, the Baltimore Abortion Fund has distributed three times the amount of funds to those who need assistance and has experienced 33% more calls to their hotline after DvJ.^19^ The National Network of Abortion Funds has also estimated their disbursement<u>s</u> to callers seeking support has more than tripled within one year after DvJ was ratified.^19^

It is not yet fully known the extent of the effects that DvJ has on patients, the healthcare system, or organizations like Planned Parenthood. To better understand the severity of DvJ on abortion-protecitve states like Colorado, there needs to be more frequent reporting in abortion incidence. Some states publish publicly-available, annual data on induced terminations; however, not all states do so. Advocating for nationwide or state policies surrounding the reporting and public dissemination of abortion incidence, at least bi-annually, would allow researchers to study the impacts of DvJ across the country more effectively.

### Limitations

The data reported by the CDPHE included state level demographic information for terminations occurring within the state from 2004 through 2023. The data and analysis were limited because the information was not linked to the patient, which made it difficult to draw conclusions about how factors like socioeconomic status are involved in the changes in abortion incidence in Colorado. Future work should collect case-specific data to estimate the association between state residence, race, and gestation week at time of abortion.

Though extensive, the CDPHE data reporting was inconsistent in reporting metrics. Demographic characteristics such as marital status, age, and race were reported monthly from 2004 through 2023. Data on state of residence was reported annually from 2004 to 2023. Inconsistencies in the frequency of reporting affected our analysis, resulting in the use of multiple different data subsets. Reported demographic data was also not inclusive of all races/ethnicities, but rather only reported (1) White non-Hispanic, (2) White Hispanic, (3) Black, and (4) Other. Our research team interpreted the *Other* category to include all excluded racial and ethnic groups. Lastly, DvJ remains a current event; there is limited data since June 2022 to study the extent of the impact on healthcare facilities or individuals.

Our study begins to understand how DvJ has resulted in changes in abortion incidence in abortion-protective states. In studying the state of Colorado, we observed a significant increase in the incidence of total terminations and attributed the change to an influx of non-resident patients from states with severely restrictive abortion legislation. Our findings suggest that as individual states across the country restrict abortion access, protective states like Colorado will continue to experience higher volumes of total terminations, and late-gestation terminations, driven by non-residents, likely impacting appointment wait times and abortion access.

## Data Availability

De-identified data were collected from the CDPHE via a data request form and the Lehigh University Human Subjects Committee determined that no Institutional Review Board approval was necessary for this study.
Data is available upon request from the Colorado Department of Public Health and Environment.

## Acknowledgements

We wish to thank Kirk Bol from the Vital Statistics Program of the Colorado Department of Public Health for his support and expertise in the data collection associated with the induced terminations of pregnancy statistics of Colorado.

## Notes

### Competing Interest Statement

The authors have declared no competing interest.

### Funding Statement

This study did not receive any funding.

